# Aortic Strain in Patients with Marfan Syndrome Long-Term After Proximal Graft Replacement Surgery – Novel Insights Enabled by Cine-CMR Biomechanical Characterization

**DOI:** 10.1101/2022.11.01.22281805

**Authors:** Robert Y. Park, Cole Latvis, Mary J. Roman, Jiwon Kim, Hannah K. Agoglia, Nicole Liberman, Pablo Villar-Calle, Raina Jain, Sheldon Liu, Lisa Rong, Maria Chiara Palumbo, Alberto Redaelli, Yadong Wang, Jay D. Humphrey, Richard B. Devereux, Giovanni Soletti, Mario F.L. Gaudino, Leonard N. Girardi, Jonathan W. Weinsaft

## Abstract

**Background:** Prosthetic graft replacement of thoracic aortic aneurysms (TAA) yields benefits but risks persist in the native aorta, especially in Marfan syndrome (MFS). Differential biomechanics between chronically grafted and native aortic regions are unknown.

**Methods:** Functional cardiac magnetic resonance (cine-CMR) imaging was performed in non-surgical MFS patients (root or ascending diameter≤4.5cm) and patients after (>1 year) proximal grafting. Analyses included mid-ascending and -descending aortic size (diameter, area) and compliance indices, including global circumferential strain (GCS), fractional area change (FAC), stiffness index, and distensibility.

**Results:** 46 MFS patients underwent cine-CMR, including 21 with chronic proximal grafts (10.5±7.3 years post-operatively). Patients with and without grafts had similar clinical and hemodynamic characteristics. Grafted and non-grafted ascending aortic size was similar between groups (p=NS), but functional parameters differed as evidenced by decrements in GCS, FAC, stiffness index, and distensibility (all p<0.001), consistent with prosthesis-associated reductions in compliance: Proximal grafts associated with decreased ascending aortic strain (B= −7.09, p<0.001) independent of age and aortic root size. Notably, native descending aortic size was larger in post-operative patients (p<0.01), paralleled by increased GCS (p<0.001) and decreased stiffness (p=0.04). In multivariate analysis, proximal grafts associated with increased descending aortic strain when controlling for ascending aortic area (B=4.19, p<0.001) or root size (B=3.14, p=0.045).

**Conclusions:** Marfan syndrome patients with chronic proximal aortic grafts manifest distinct vessel wall biomechanics in grafted and native regions that differ from non-surgical comparators, including decreased strain (a marker of reduced compliance) within grafted territories and increased strain in native aortic regions distal to grafts.

## Introduction

Surgical prosthetic graft replacement is a therapeutic cornerstone for managing thoracic aortic aneurysms (TAA).^1,2^ Prosthetic graft surgery eliminates risk for dissection in graft-replaced regions, but risk for long-term complications in the distal (native) aorta persists: Aortic events following proximal grafting are markedly increased in patients with Marfan syndrome, a group that typically undergoes prosthetic graft surgery at lower thresholds, higher frequency, and younger age: For example, in prior multicenter research, two thirds (68%) of dissections in Marfan patients occurred after proximal graft surgery.^3^ Similarly, other studies have reported Marfan patients to be at increased risk for distal aortic aneurysm, re-operation, and dissection after proximal grafting.^4,5^ Given the seriousness of such events, further research into mechanism of adverse remodeling after proximal grafting is of substantial importance.

One reason for aortic events after TAA replacement surgery may stem from biomechanical properties of grafts: Prosthetic grafts are stiffer than native aortic tissue and tubular in shape, thus providing a nidus for energy propagation to the distal aorta that increases stress on native tissue. Consistent with this, prior studies from our group using echocardiography showed that proximal grafts increase distal aortic wall excursions (strain) intra-^6^ and post-operatively.^7^ Our cardiac MRI (CMR) studies demonstrated further that proximal grafting produces short-term increases in both ascending and descending aortic flow velocity, flow-induced wall shear stress, and pressure-induced wall strain.^8^ While these studies support the notion that grafts alter biomechanics, findings have been derived from mixed diagnostic cohorts, focused exclusively on the descending aorta, and/or entailed short term follow-up: Aortic biomechanics - including ascending and descending aortic vessel wall compliance - in patients long-term after proximal graft implantation are unknown.

This study used CMR to assess thoracic aortic biomechanics in non-surgical and post-surgical Marfan syndrome patients well after (>1 year) proximal aortic grafting. To do so, functional CMR was analyzed in the ascending and descending aorta to quantify vessel wall displacement (strain) in graft replaced and distal native regions. The goal was to test the hypothesis that Marfan patients with chronic grafts manifest differences in aortic vessel wall compliance, both within graft replaced regions and in downstream native aortic territories.

## Methods

### Study Population

The population comprised TAA patients with clinically documented Marfan syndrome undergoing elective surgical prosthetic graft replacement of the ascending thoracic aorta, as well as Marfan patients with aortic root and ascending aortic diameters below thresholds for cutoffs for consideration of elective graft replacement (root or ascending aortic diameter ≤ 4.5cm).^1^ To assess the impact of grafts on aortic biomechanics in a uniform cohort with minimal confounders, patients with pre-operative aortic dissection were excluded from the study. Patients were included in the study based on the above clinical criteria, and availability of cine-CMR images with which to quantify aortic deformation (strain).

Aortic surgery was performed between 1994 and 2019 using conventional prosthetic (polyethylene terephthalate; Dacron) grafts. CMR exams were retrieved from data archives and analyzed (blinded to demographic and cardiac structural/functional indices) to quantify aortic functional indices within native and grafted regions. This study was conducted with approval of the Weill Cornell Institutional Review Board, which approved use of pre-existing data as was analyzed for study-related research purposes.

### Image Acquisition

CMR was performed for clinical indications using commercial (1.5 [24%] or 3.0 [76%] Tesla) scanners: Study-related data were derived from cine-CMR, which was acquired using a steady-state free precession pulse sequence (typical parameters: flip angle 60°, TR 3.5msec, TE 1.5 msec, slice thickness 6.0mm). Routine cine-CMR images were used for study analyses; acquisition was not tailored for research purposes. All exams included short axis views of the mid-descending aorta; adjunctive imaging of the ascending aorta was available in 83.0% (n=38/46) of cases.

### Image Analysis

#### Aortic Compliance and Geometry

Post-processing and aortic strain analysis were performed offline using dedicated software (Precession [HeartIT, Raleigh, NC]). Briefly, after manually tracing luminal boundaries of the aortic wall at an initial time, automatic vessel wall tracking was used to measure motion throughout subsequent frames over the cardiac cycle: If computer-generated analysis was deemed suboptimal (e.g. discordant with visualized aortic contours), seed points were manually adjusted to optimize aortic wall vessel tracking. Aortic frames corresponding to maximum and minimum area (respectively labeled as end-systolic area and end-diastolic area) were used for circumferential strain computation: GCS= (Cs-Cd)/Cd x 100, where Cs and Cd are circumference, respectively, of the systolic and diastolic area.

Ancillary parameters used to assess aortic compliance included time to peak strain (calculated as time interval between aortic end-diastole and end-systole), distensibility (ESA – EDA)/(EDA x [SBP-DBP]), where SBP and DBP denote systolic and diastolic blood pressure respectively, and EDA and ESA denote the cross sectional luminal area of the descending aorta at end systole and end diastole, and fractional area change ([ESA-EDA]/ESA) – each of which were measured concordant with our prior research.^8,9^ Aortic stiffness index (β) was calculated using an established formula (ln (SBP / DBP)/([ESA – EDA]/EDA).^10^ Aortic diameter was measured at landmarks throughout the aorta; root dimensions were used for z-score calculations in non-surgical patients using normative reference data.^11^ Aortic area was measured in locations corresponding to strain analyses (mid ascending, descending aorta).

#### Left Ventricular Function and Remodeling

Left ventricular (LV) indices were available in 83% of patients and were quantified were quantified via planimetry of end-diastolic and end-systolic chamber volumes, which yielded LV ejection fraction and stroke volume. LV mass was quantified by planimetry of LV end-diastolic endocardial and epicardial borders; papillary muscles and trabeculae were excluded from chamber volumes (included in myocardial mass). LV analyses were performed via a dedicated study investigator (JWW) for whom high reproducibility has been previously reported.^12^

### Clinical and Hemodynamic Assessments

Clinical indices, including surgical and demographic parameters as well as medication regimen, were extracted via electronic medical record review. Post-operative intervals were calculated in relation to CMR. Blood pressure was determined via brachial measurements. Heart rate was attained at the time of CMR.

### Statistical Analyses

Comparisons between groups were made using Student’s t test (expressed as mean value ± standard deviation) for continuous variables. Categorical variables were compared using Chi-square or, when fewer than 5 expected outcomes per cell, Fisher’s exact test. Univariable and multivariable linear regressions analyses were used to assess associations between indices, including strain in relation to prosthetic graft implantation. Receiver operating curve (ROC) analysis was used to evaluate diagnostic performance of CMR strain parameters for identification of patients with ascending aortic grafts. Two-sided p<0.05 was considered indicative of statistical significance. Statistical calculations were performed using SPSS 27.0 (SPSS Inc. [Chicago, IL]).

## Results

### Population Characteristics

The study group comprised 46 patients with Marfan syndrome, 21 of whom underwent elective replacement of a TAA with a prosthetic graft and 25 non-surgical patients (maximal root or ascending aorta ≤4.5cm [root diameter 4.1±0.3cm; z-score 2.9±1.2]): All post-operative patients had graft implantation >1 year prior to CMR (mean post-operative interval 10.5±7.3 years); 76% (n=16) underwent valve sparing root repair and 24% (n=5) had concomitant aortic valve replacement.

**Table 1** reports characteristics of the study population, including comparisons between post-operative and non-surgical cohorts: As shown, groups did not differ significantly with respect to clinical characteristics, hemodynamic indices, or medication regimen, although post-operative patients tended to be older (p=0.06). Similarly, as shown in **Table 2**, LV geometry and functional indices (quantifiable on CMR in 83.0% of patients) were similar between groups, including near equivalent LVEF and stroke volume (both p=NS).

**Table 1.**
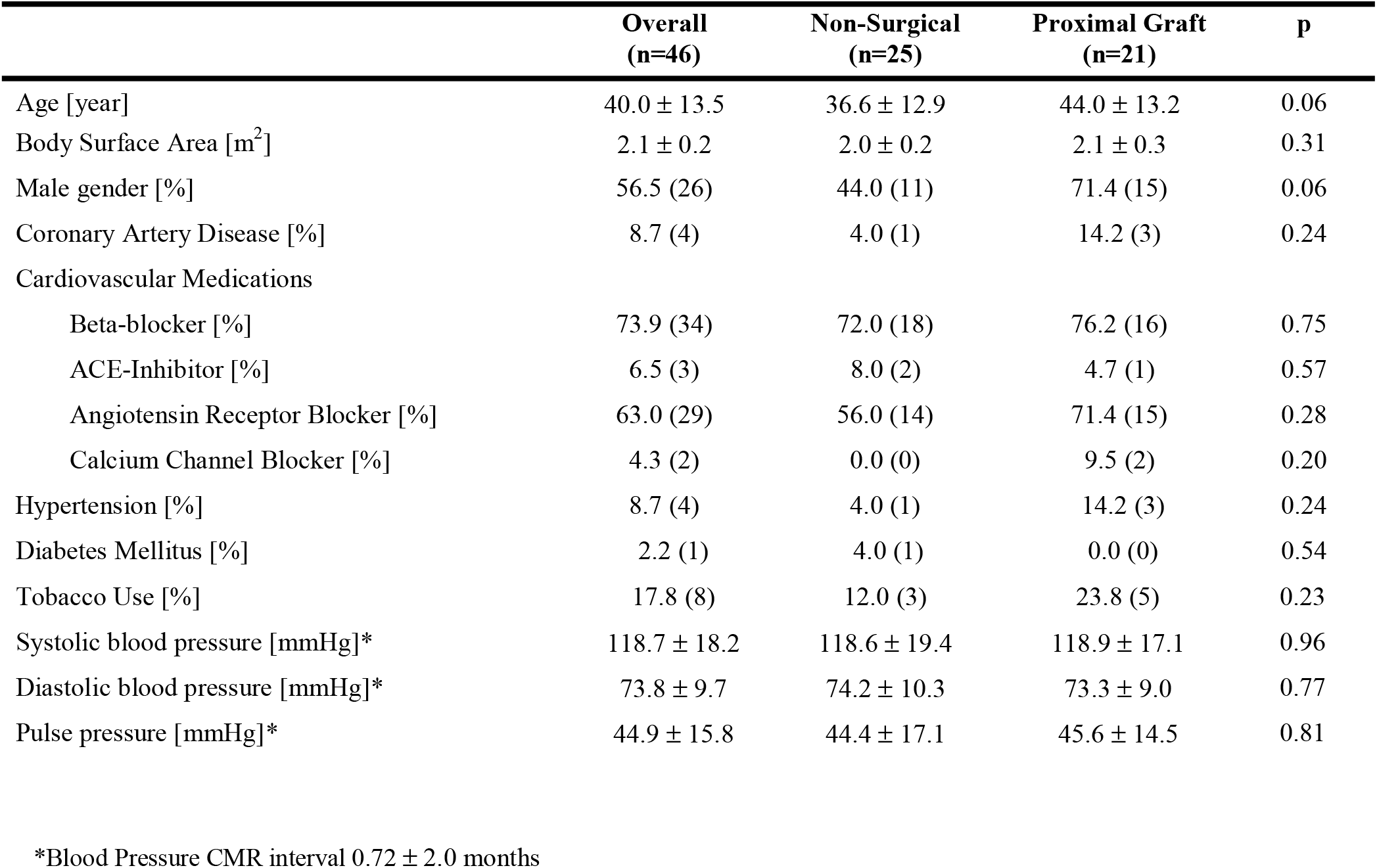
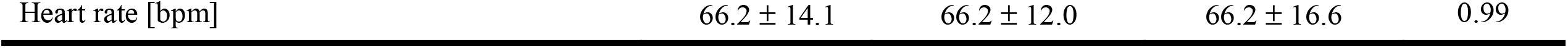
Clinical Characteristics.

**Table 2.** Left Ventricular Function and Geometry*.

|  | Overall | Non-Surgical | Proximal Graft | p |
| --- | --- | --- | --- | --- |
| Imaging Variables |  |  |  |  |
| LV End-Diastolic Volume [ml/m<sup>2</sup>] | 85.6 ± 24.3 | 83.8 ± 22.1 | 87.8 ± 27.3 | 0.62 |
| LV End-Systolic Volume [ml/m<sup>2</sup>] | 32.1 ± 17.3 | 30.5 ± 11.2 | 34.1 ± 23.0 | 0.53 |
| LV Stroke Volume [ml] | 111.7 ± 28.1 | 109.1 ± 26.4 | 114.8 ± 30.7 | 0.55 |
| LV Cardiac Output [l/min] | 6.6 ± 1.9 | 6.4 ± 1.4 | 6.9 ± 2.4 | 0.44 |
| LV Ejection Fraction [%] | 63.7 ± 9.1 | 64.1 ± 6.8 | 63.2 ± 11.5 | 0.77 |
| LV Mass [g/m<sup>2</sup>] | 66.5 ± 15.8 | 62.0 ± 10.2 | 72.4 ± 20.0 | 0.08 |

\*Available in 83% (38/46) of patients

### Aortic Dynamic and Geometric Indices

**Table 3** reports strain-related variables together with conventional indices of aortic size. In the ascending aorta (post-operatively grafted region), results demonstrate over a 2-fold decrease in GCS, FAC, stiffness index (β) and distensibility among post-operative, compared to non-surgical, Marfan patients (all p<0.001): Consistent with this, ROC analysis demonstrated proximal aortic strain indices to yield high overall performance for differentiating between non-surgical and grafted patients (GCS AUC: 0.986 [CI 0.956, 1.000], |FAC: 0.997 [0.988, 1.000]; both p<0.001), corresponding to high sensitivity and specificity (GCS cutoff-6.13: 0.952, 1.000 | FAC cutoff-8.62: 0.952, 1.000). Strain-related differences between groups were paralleled by slightly larger aortic size among non-surgical patients, as evidenced by increased root diameter (p<0.001) and a trend towards increased ascending aortic (end-systolic) area (p=0.08).

**Table 3.** Cine-CMR quantified aortic geometry and vessel wall dynamics.

|  | Ascending Aorta* |  |  | Distal Aorta |  |  |
| --- | --- | --- | --- | --- | --- | --- |
|  | Non-Surgical<br>(n=21) | Proximal Graft<br>(n=17) | p | Non-Surgical<br>(n=25) | Proximal Graft<br>(n=21) | p |
| **Geometric Indices** |  |  |  |  |  |  |
| End-diastolic Area [cm<sup>2</sup>] | 6.2 ± 2.0 | 6.3 ± 1.8 | 0.96 | 3.7 ± 1.1 | 4.8 ± 1.5 | **0.004** |
| End-systolic Area [cm<sup>2</sup>] | 7.7 ± 2.2 | 6.5 ± 1.8 | 0.08 | 4.3 ± 1.1 | 5.9 ± 1.6 | **<0.001** |
| Maximal Diameter [cm] | 3.3 ± 1.2 | 2.9 ± 0.4 | 0.19 | 2.3 ± 0.2 | 2.7 ± 0.4 | **<0.001** |
| Root Diameter [cm] | 4.1 ± 0.3 | 3.2 ± 0.5 | **<0.001** |  |  |  |
| Arch Diameter [cm] |  |  |  | 2.5 ± 0.3 | 2.7 ± 0.4 | **0.01** |
| **Dynamic Indices** |  |  |  |  |  |  |
| Circumferential Strain [%] | 10.9 ± 3.5 | 3.8 ± 1.2 | **<0.001** | 6.0 ± 2.3 | 10.3 ± 4.4 | **<0.001** |
| Fractional Area Change [%] | 19.9 ± 6.9 | 3.6 ± 1.7 | **<0.001** | 13.9 ± 8.0 | 18.6 ± 8.1 | 0.052 |
| Stiffness Index (β) | 2.2 ± 1.4 | 15.5 ± 7.3 | **<0.001** | 4.8 ± 4.5 | 2.7 ± 1.7 | **0.04** |
| Distensibility [10<sup>-3</sup>mmHg<sup>-1</sup>] | 6.8 ± 3.6 | 0.9 ± 0.4 | **<0.001** | 4.6 ± 3.8 | 6.0 ± 3.9 | 0.22 |
| Time-to-Peak Strain [ms] | 297.7 ± 72.3 | 283.9 ± 89.7 | 0.60 | 258.0 ± 64.8 | 309.4 ± 180.8 | 0.19 |

\*Available in 83% (38/46) of patients

**Table 3** also demonstrates that proximal grafting was accompanied by alterations in descending thoracic aortic vessel wall dynamics. GCS was higher among post-operative patients (p<0.001), paralleled by decreased stiffness (p=0.04). Increased vessel wall strain occurred despite larger descending aortic size, as evidenced by increased area and diameter (both p<0.01). In sub-group analysis of patients who underwent graft implantation with adjunctive valve sparing root repair but no valve replacement (n=16), descending aortic GCS (11.7 ± 4.0 vs. 6.0 ± 2.3, p <0.001) and FAC (20.0 ± 8.1 vs. 13.9 ± 8.0%, p=0.03) were significantly higher compared to non-surgical patients, paralleled by decreased stiffness index (2.4 ± 1.5 vs. 4.8 ± 4.5, p=0.02).

**Figure 1** provides representative examples of aortic strain in a non-surgical and chronically grafted patient (post-operative interval 3.25 years), demonstrating diminished compliance within grafted territories paralleled by augmentation in the descending (native) aorta.

**Figure 1:**
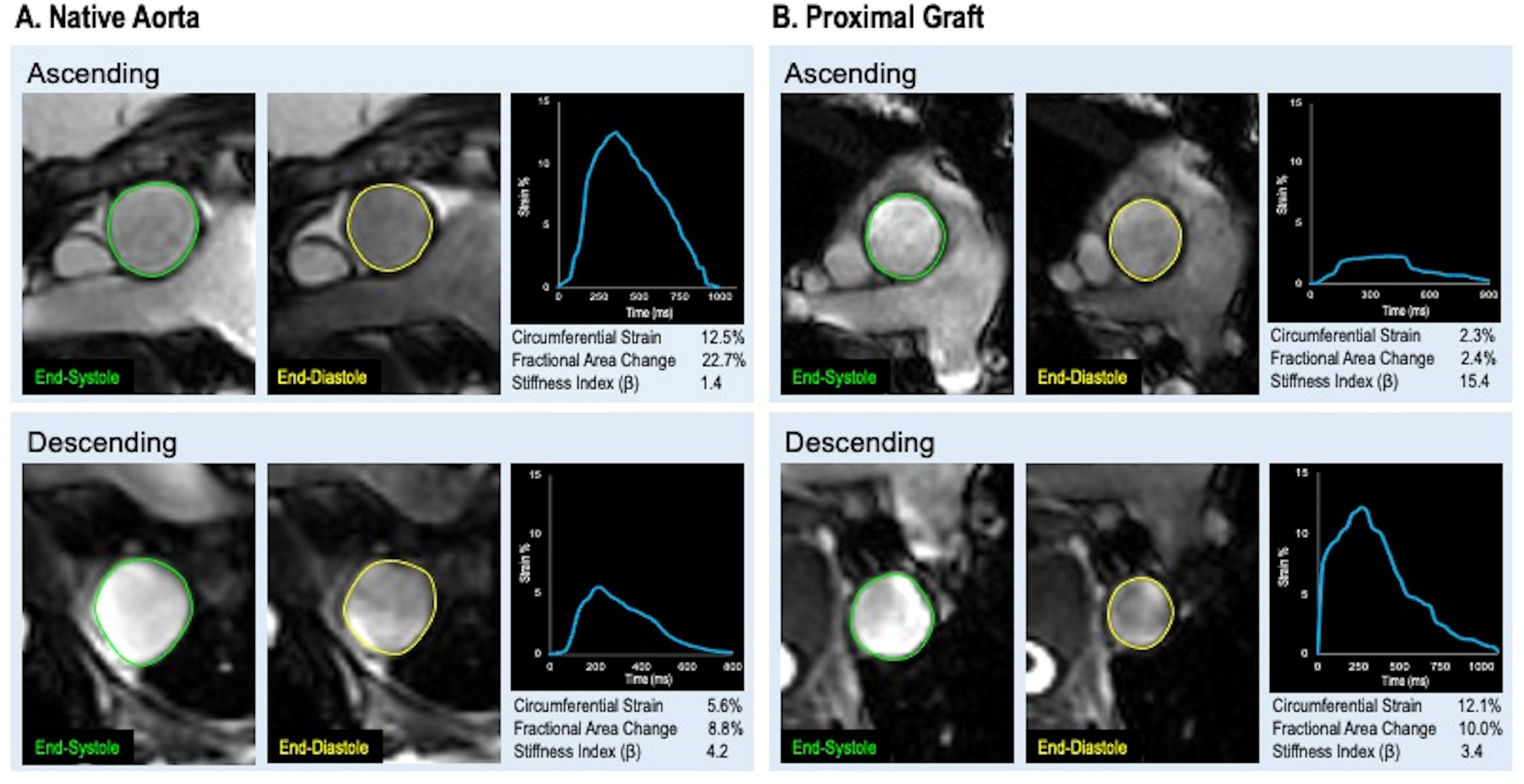
Representative examples of global circumferential (GCS) strain analysis in the mid ascending (top) and mid descending (bottom) aorta among Marfan syndrome patients with (left) and without (right) proximal aortic grafts. Note decreased aortic circumferential strain (reduced compliance) within grafts as compared to compared to the native ascending aorta, paralleled by increased strain in regions distal to grafts. Global circumferential strain is represented by blue curves (right); corresponding end-systolic and end-diastolic border delineation are represented by green and yellow circles.

## Markers of Aortic Strain

### Ascending (Grafted) Aorta

**Table 4** reports regression analyses of clinical and imaging indices in relation to ascending aortic strain (GCS). As shown, prosthetic graft surgery associated with decreased strain in the ascending aorta whereas aortic root diameter (as was increased among the non-surgical cohort) associated with increased strain (both p<0.001): In multivariate analysis (**4B**), proximal graft implantation (B = −7.09 [CI −9.96 – −4.21], p<0.001) was independently associated with decreased ascending aortic GCS when controlling for age and aortic root size (both p=NS).

**Table 4.**
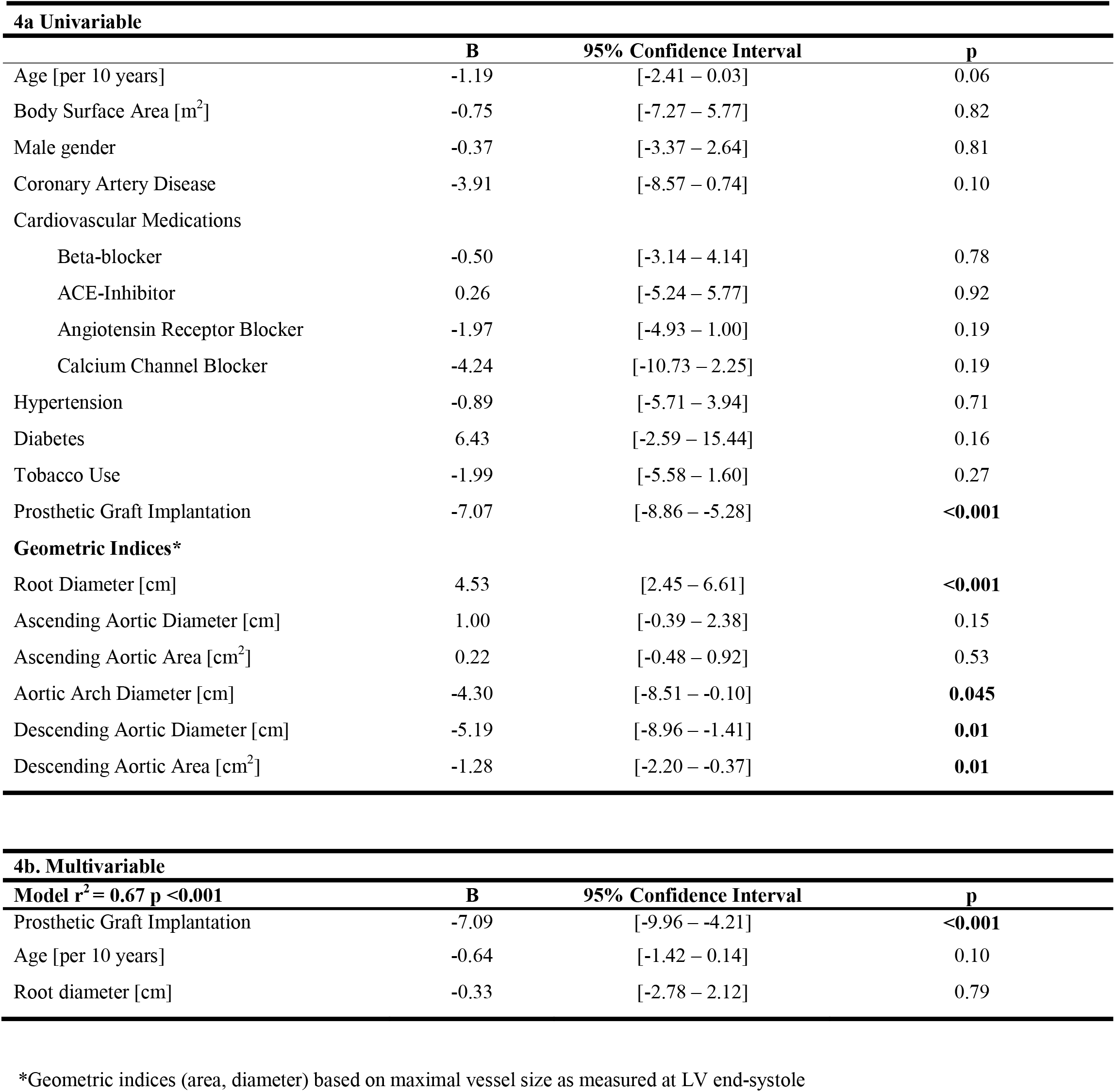
Regression Analysis for Ascending Aortic Strain.

### Descending (Native) Aorta

**Table 5** reports corresponding regression analyses in relation to descending aortic strain (GCS). Results demonstrate that proximal grafting associated with increased descending aortic GCS whereas root diameter and ascending aortic area (as were increased in non-surgical patients) associated with decreased GCS (both p<0.05). In multivariable analysis (**5B**), proximal grafting independently associated with increased descending aortic GCS when controlling for ascending aortic area (B = 4.19 [CI 2.10 – 6.29], p<0.001). Results were similar in multivariable analysis inclusive of prosthetic grafting and aortic root size (**5C**) for which proximal grafts were also independently associated with increased GCS in the native descending aorta (B=3.14 [CI 0.07 – 6.20], p=0.045).

**Table 5.**
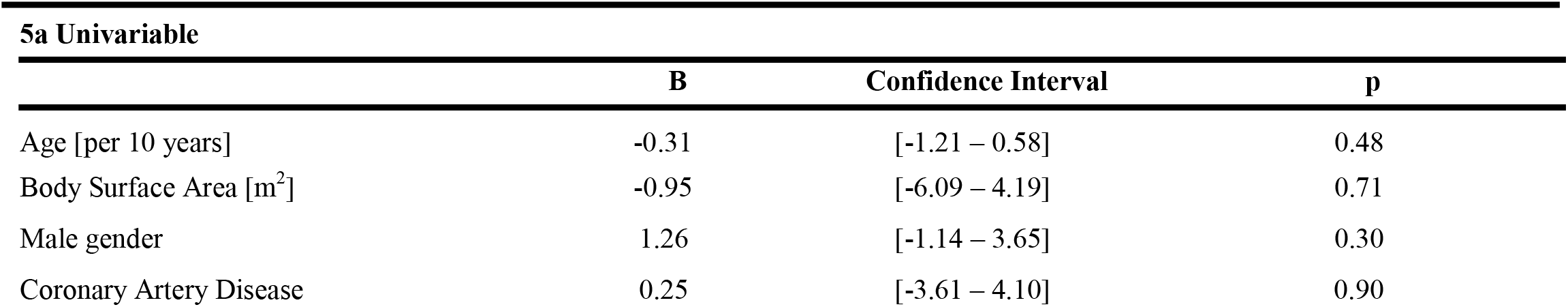
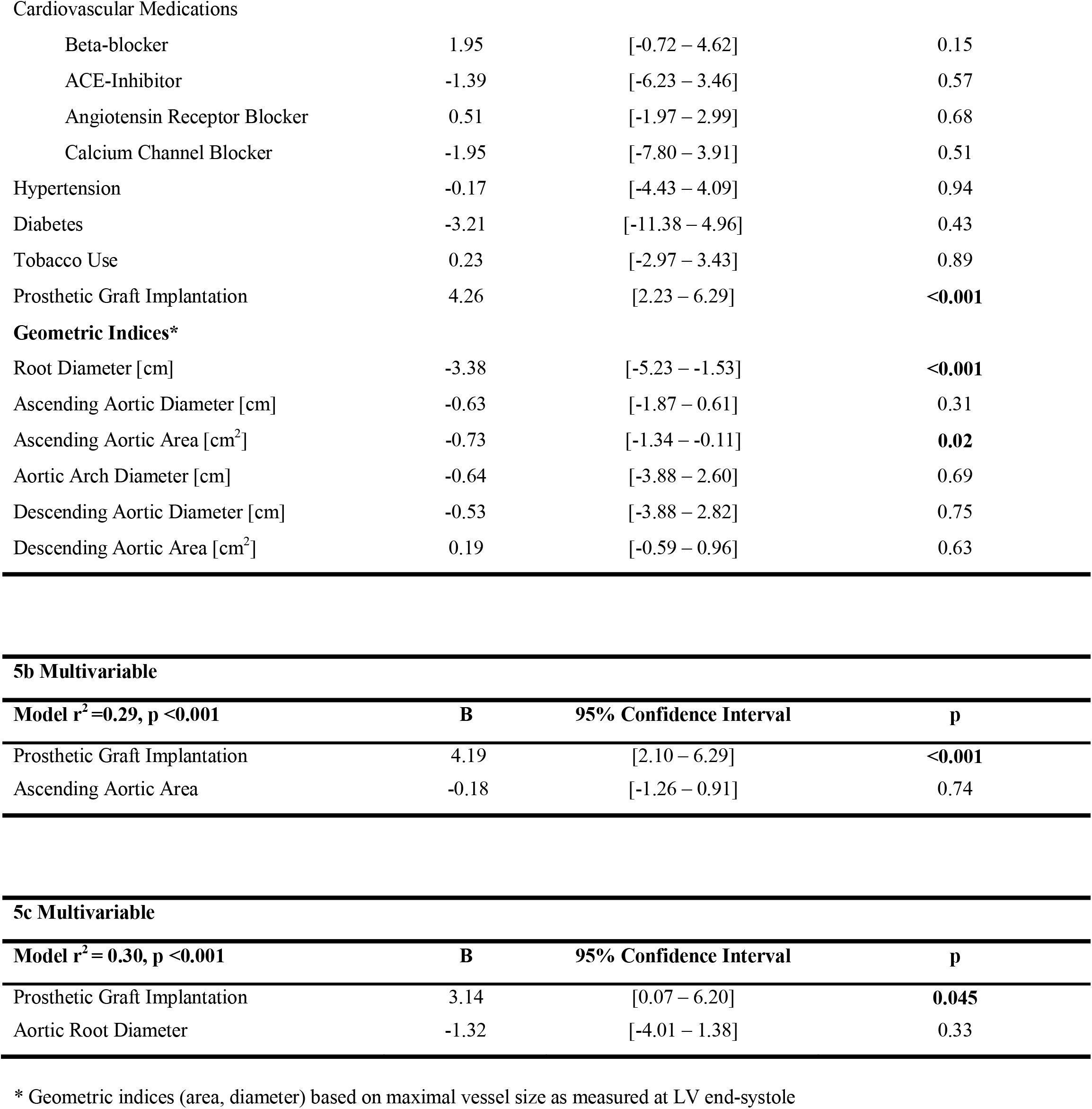
Regression Analysis for Descending Aortic Strain.

## Discussion

This study provides multiple new insights regarding differential aortic biomechanics long-term after proximal aortic graft repair of TAA in patients with Marfan syndrome: First, among a cross-sectional cohort of patients with and without proximal grafting, aortic compliance differed in relation to surgical status, as evidenced by decreased aortic strain within grafted regions and associated increased aortic strain within the distal native aorta: Differences were evident chronically after graft implantation (mean post-operative duration 10.5±7.3 years). Second, strain-related differences between grafted and native (ascending) aortic territories were evident despite similar ascending aortic size, corresponding to high overall diagnostic performance of strain and FAC quantification to differentiate between post-operative and non-surgical patients (AUC=0.986 and 0.997 respectively, both p<0.001). Third, proximal graft implantation related strongly related to differential strain in surgically-replaced and native aortic territories, as evidenced by independent associations between graft status and both decreased strain in the ascending aorta (B = −7.09 [CI −9.96 – −4.21], p<0.001) and increased strain in the descending aorta in separate multivariable analyses controlling for aortic area (B=4.19, p<0.001) or root size (B=3.14, p=0.045).

Whereas this is the first study that we are of aware of to examine ascending and descending aortic compliance patterns in patients long-term after proximal graft implantation, our findings add literature by our group and others demonstrating that differences in mechanical properties between current prosthetic grafts and the native aorta alter aortic wall biomechanics in regions distal to surgically treated territories. This premise is supported by both imaging^6–8^ and computational modeling studies.^13–15^ In clinical studies, graft-induced changes in the distal aorta have been shown to be greatest in patients with genetically triggered TAA – in whom we^16^ and others^17–19^ have shown aortic stiffness to be greatest. Increased stiffness in native regions distal to grafts can augment energy propagation to the descending aorta. This is important as altered hemodynamics can contribute to internal elastic lamina fragmentation and vessel tortuosity, which have been linked to dissection.^20^ These concepts support a paradigm in which proximal graft implantation augments distal energy propagation and vessel wall strain, providing a nidus for adverse remodeling in susceptible patients such as those with Marfan syndrome.

Applied clinically, our findings support the notion that aortic strain and related compliance indices – as can be derived from post-processing of routine clinical datasets widely used for functional MRI, may provide value for identification of aortic grafts. It is also possible that strain provides prognostic value for anticipating aortic events after graft replacement surgery. In our study, differences in aortic strain occurred despite equivalent ascending aortic size and corresponded to high performance for differentiating surgical and non-surgical patients. Other groups have demonstrated that aortic strain can differ between aortopathy types,^16–19^ and can vary in relation to altered hemodynamics secondary to aortic valve disease,^21^ renal dysfunction,^22^ and systemic hypertension.^23^ Taken together, these data support the notion that biomechanical strain-based analyses can provide insights of clinical and mechanistic value that are additive to those of conventional geometric aortic assessments.

Several limitations should be recognized. First, findings were derived from pre-existing clinical cine-CMR images rather than tailored pulse sequences to quantify flow, wall thickness, and tissue characterization indices. While our approach supports the notion that strain analysis enables novel mechanistic insights to be derived from routine imaging datasets, it is uncertain whether changes in aortic vessel wall biomechanics are paralleled by differences in flow or other additional indices. It should also be noted that blood pressure was measured via peripheral (rather than central methods) and was not obtained concomitantly with CMR, and that other potential confounders (including aortic regurgitation) were not quantified. Second, whereas post-operative follow-up duration was long, imaging was acquired at a single time point that varied between patients, thus prohibiting analyses as to whether native distal aortic remodeling is progressive after graft implantation. Third, whereas this study focused on aortic strain after proximal grafting, it is unknown whether increased strain corresponds to augmented clinical event risk. Finally, the present approach is unable to examine possible effects of increased wall strain on distal aortic wall microstructure and its potential vulnerability to dissection or rupture. Such would require associated histopathology, which is not available. Studies in mouse models of Marfan syndrome reveal histo-mechanical changes in the descending thoracic aorta that only are different in extent, though not type, from the ascending aorta^24,25^ suggesting that proximal graft-related increases in mechanical loading on the distal segment could exacerbate the underlying disease in this segment. Further research will be needed to examine this possibility.

In conclusion, results of this study demonstrates that Marfan patients undergoing prosthetic graft replacement surgery demonstrate long-term differences in aortic compliance as compared to non-surgical patients, including increased strain in distal native aortic territories. Future prospective studies are warranted to test whether post-operative aortic remodeling is progressive and/or varies in relation to surgical approaches, if increased aortic strain predicts clinical event risk after proximal graft surgery, and if differences in distal aortic biomechanics and clinical events after proximal graft repair are driven by graft effects or by native aortic phenotype.

## Data Availability

All data produced in the present study are available upon reasonable request to the authors

